# Severe COVID-19 is characterised by inflammation and immature myeloid cells early in disease progression

**DOI:** 10.1101/2021.09.01.21262953

**Authors:** Liam Townsend, Adam H Dyer, Aifric Naughton, Sultan Imangaliyev, Jean Dunne, Rachel Kiersey, Dean Holden, Aoife Mooney, Deirdre Leavy, Katie Ridge, Jamie Sugrue, Mubarak Aldoseri, Jo Hannah Kelliher, Martina Hennessy, Declan Byrne, Paul Browne, Christopher L Bacon, Catriona Doyle, Ruth O’Riordan, Anne-Marie McLaughlin, Ciaran Bannan, Ignacio Martin-Loeches, Arthur White, Rachel M McLoughlin, Colm Bergin, Nollaig M Bourke, Cliona O’Farrelly, Niall Conlon, Clíona Ní Cheallaigh

## Abstract

SARS-CoV-2 infection causes a wide spectrum of disease severity. Immune changes associated with severe disease include pro-inflammatory cytokine production and expansion of immature myeloid populations. The relative importance of the immunological changes in driving progression to severe disease remain poorly understood.

We aimed to identify and rank clinical and immunological features associated with progression to severe COVID-19. We sought to use tests available in an on-site diagnostic hospital laboratory to identify an immunological signature for severe disease development which could be detected prior to peak severity thereby allowing initiation of therapeutic interventions. We used univariate and multivariate analysis, including unbiased machine learning, to investigate the relationships between clinical and demographic characteristics, inflammatory markers, and leukocyte immunophenotypes with progression to severe disease in 108 patients and to rank these in importance. A combination of four features (elevated levels of interleukin-6 and C-reactive protein, coupled with reduced monocyte HLA-DR expression and reduced neutrophil CD10 expression), were strongly predictive of severe disease with an average prediction score of 0.925.

**Highlights:**

- Severe COVID-19 can be predicted by a combination of emergency myelopoiesis (CD10-neutrophils and HLA DR-monocytes) and inflammation (raised IL-6 and CRP)
- These changes can be identified from tests carried out prior to peak illness severity in a diagnostic laboratory
- This predictive model was derived from a cohort of patients with a wide range of ages, frailty and COVID-19 severity

**Graphical Abstract:** 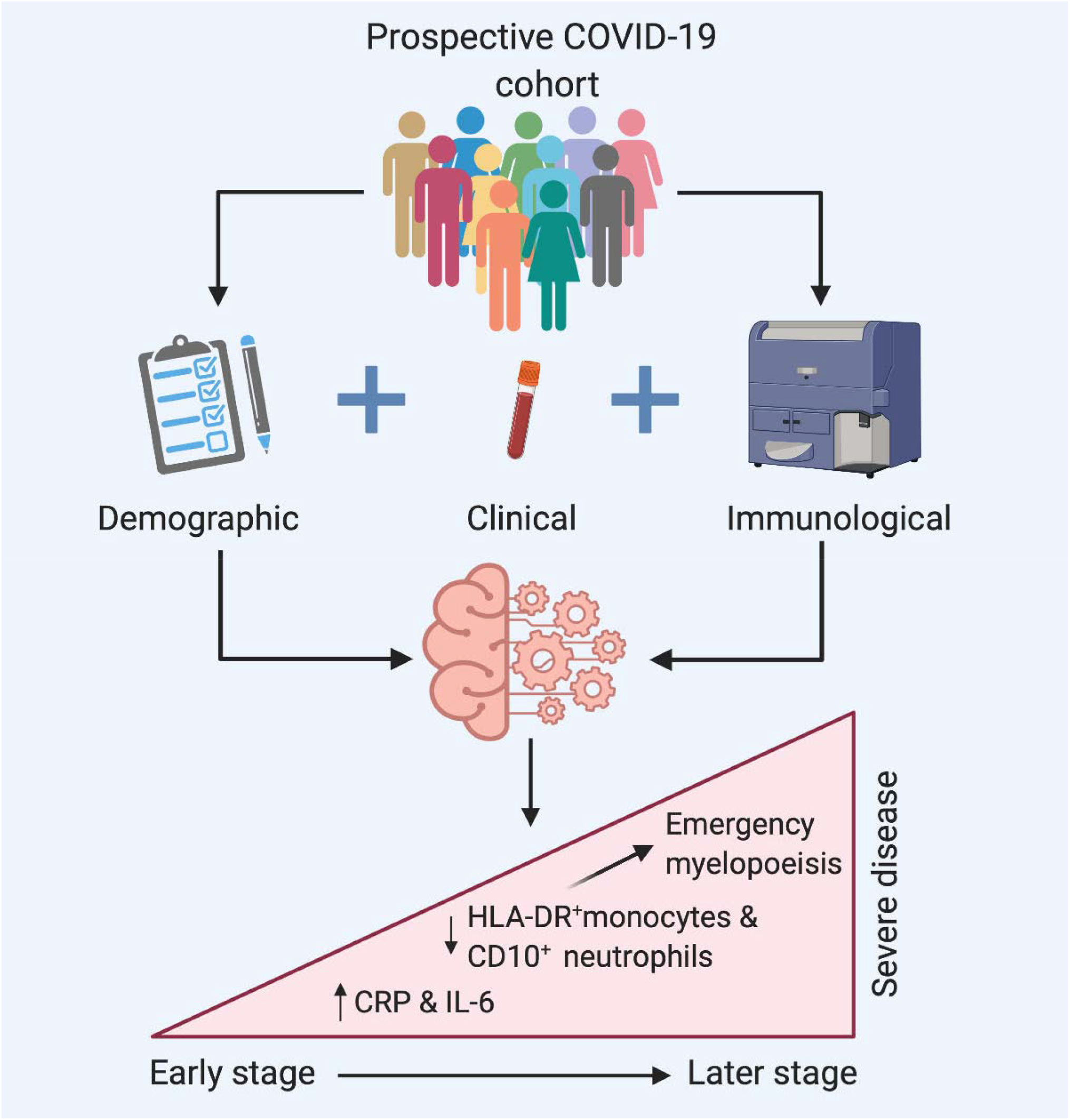

## Introduction

COVID-19, caused by infection with the SARS-CoV-2 virus, is responsible for the current global pandemic.^1^ The significant morbidity and mortality associated with this infection has placed unprecedented pressures on healthcare systems worldwide.^2^ The clinical course of SARS-CoV-2 infection is remarkably variable. COVID-19 appears to have a biphasic pattern of illness.^3-5^ The initial phase, due to viral replication and the initial immune response, is characterised by elevation in inflammatory cytokines and influx of monocytes and T lymphocytes into the lungs.^6-10^ An immune-mediated phase develops in approximately 20% of individuals approximately 6-10 days post symptom onset.^11^ This phase is characterised by further elevation of inflammatory markers, increase in D-dimer levels, shortness of breath, and lung infiltrates.^12,13^ In approximately 5% of cases, progressive difficulty in breathing occurs, resulting in ARDS and requirement for mechanical ventilation.^14^

Multiple recent studies have described diverse immune mechanisms underlying COVID-19 pathogenesis. Most have looked at individual associations between immune variables and disease severity and identified alterations in cytokine levels and within immune cell populations.^15-20^ However, the pathophysiology of COVID-19 results from complex interactions across multiple parameters. Looking at individual associations between immune variables and disease in isolation using simple univariate statistical hypothesis testing cannot assess such interactions. Integration of multiple immunological and clinical parameters using multivariate testing is required.^21^ In addition, interventions carried out in the care of patients with severe COVID-19 including intubation and mechanical ventilation are also likely to cause alterations in immune parameters, and it is therefore important to study immune mechanisms early in the course of disease. Recent reports have highlighted severity-associated changes in myeloid and lymphocyte subsets but the relative importance of such changes in relation to other important demographic and clinical risk factors that can influence trajectories of disease progression is unknown.^19, 20^ Finally, the rapid and early identification of those who will develop severe disease is essential for the prompt administration of therapies. Dexamethasone has been shown to reduce mortality in patients requiring oxygen therapy.^22^ However, administering this in a timely fashion has proven difficult.^23^ There is an unmet need in for accurate immunological predictors of severe disease which can be used in a clinical setting.

In this study, we describe early clinical and immunological features in one hundred and eight patients with COVID-19, of whom thirty-nine progressed to severe disease. All results were generated on the same day as the sample was obtained. We objectively ranked these features as independent predictors for COVID-19 severity and confirmed the important role for emergency myelopoiesis as demonstrated by increased proportions of immature neutrophils and HLA-DR-monocytes in combination with elevated levels of IL-6 and CRP, acting as proof of concept that these markers can be reliably and rapidly used in routine clinical practice to identify patients who will progress to severe disease and allow for the timely administration of therapeutics.

## Results

### Participant characteristics

One-hundred and eight participants were recruited (42/108 (38.9%) female). Baseline characteristics are shown in **Table 1**. Seventeen had mild disease, 52 had moderate disease (of whom 30 required supplemental oxygen) and 39 had severe disease (**Table 2**). Of those with severe disease, 26/39 were admitted to ICU for intensive monitoring and mechanical ventilation and an additional 13/39 patients were assessed for ICU admission for mechanical ventilation but were not admitted to ICU due to high likelihood of non-survival. Ten patients with severe COVID-19 died (2/26 admitted to ICU and 8/13 assessed but not admitted to ICU). The median inpatient stay for those with moderate/severe disease was 11.5 days (range 2 – 108). The median length of stay in ICU was 12 days (range 1 – 39). Disease characteristics are shown in **Table 2**, while baseline laboratory parameters are shown in **Table 3**. As shown in **Supplemental Table 2**, clinical blood tests were conducted at a median of six days post symptom onset and, for those requiring supplemental oxygen, a median of 1.5 days prior to peak oxygen requirement, which was deemed to be a marker of peak respiratory illness.

**Table 1:** Baseline Demographics.

| Characteristic |  |
| --- | --- |
| Total number of patients | 108 |
| Admitted to hospital, number of patients (percentage of total) | 91 (84%) |
| Mild disease | 17 (16%) |
| Moderate disease | 52 (48%) |
| Severe disease | 39 (36%) |
| Date of first SARS-CoV-2 positive sample, range | 10/3/2020-2/5/2020 |
| Interval between symptom onset and peak oxygen requirement, median (range), days | 8 (2-15) |
| Requirement for supplemental oxygen, number of patients (% of total) | 65 (60%) |
| Peak CRP, median (range), mg/L | 94 (1-407) |
| Chest X-ray changes, number of patients (% of total) |  |
| *No CXR done (outpatients)* | 17 (16%) |
| *No CXR changes:* | 30 (28%) |
| *CXR changes consistent with COVID-19:* | 49 (45%) |
| *Other/Indeterminate CXR changes:* | 12 (11%) |
| Requirement for ICU (% of total) |  |
| *Not required:* | 69 (64%) |
| *Admitted to ICU:* | 26 (24%) |
| *Assessed for ICU, deemed unsuitable:* | 13 (12%) |
| Outcome |  |
| *Full recovery* | 82 (76%) |
| *Death* | 10 (9%) |
| *Residual morbidity (including decreased independence)* | 9 (8%) |
| *Outcome still unclear* | 7 (6%) |

**Table 2:** Disease Characteristics.

|  |  |
| --- | --- |
| Characteristic |  |
| Total number of patients | 108 |
| Admitted to hospital, number of patients (percentage of total) | 91 (84%) |
| Mild disease | 17 (16%) |
| Moderate disease | 51 (47%) |
| Severe disease | 39 (36%) |
| Date of first SARS-CoV-2 positive sample, range | 10/3/2020-2/5/2020 |
| Interval between symptom onset and peak oxygen requirement, median (standard deviation), days | 8 (6.3) |
| Requirement for supplemental oxygen, number of patients (percentage of total) | 65 (60%) |
| Peak CRP, median (standard deviation), mg/L | 94 (114) |
| Chest X-ray changes, number of patients (percentage of total) |  |
| *No CXR done (outpatients)* | 17 (16) |
| *No CXR changes:* | 30 (28) |
| *CXR changes consistent with COVID-19:* | 49 (45) |
| *Other/Indeterminate CXR changes:* | 12 (11) |
| Requirement for ICU (percentage of total) | 69 (64)26 (24) |
| *Not required:* | 13 (12) |
| *Admitted to ICU:* |  |
| *Assessed for ICU, deemed unsuitable:* |  |
| Outcome (percentage of total) |  |
| *Full recovery* | 82 (76%) |
| *Death* | 10 (9%) |
| *Residual morbidity (including decreased independence)* | 9 (8%) |
| *Outcome still unclear* | 7 (6%) |

**Table 3:**
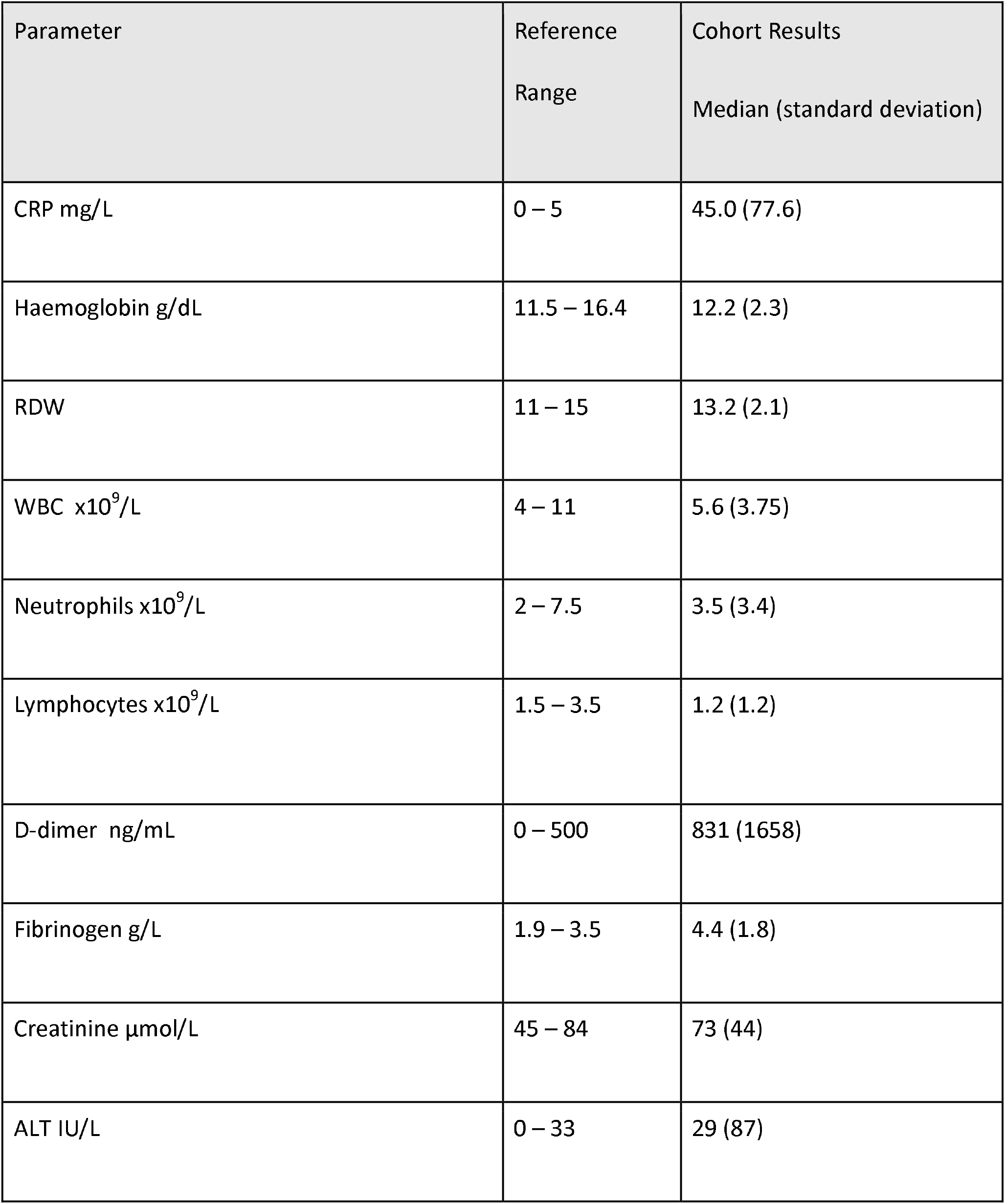
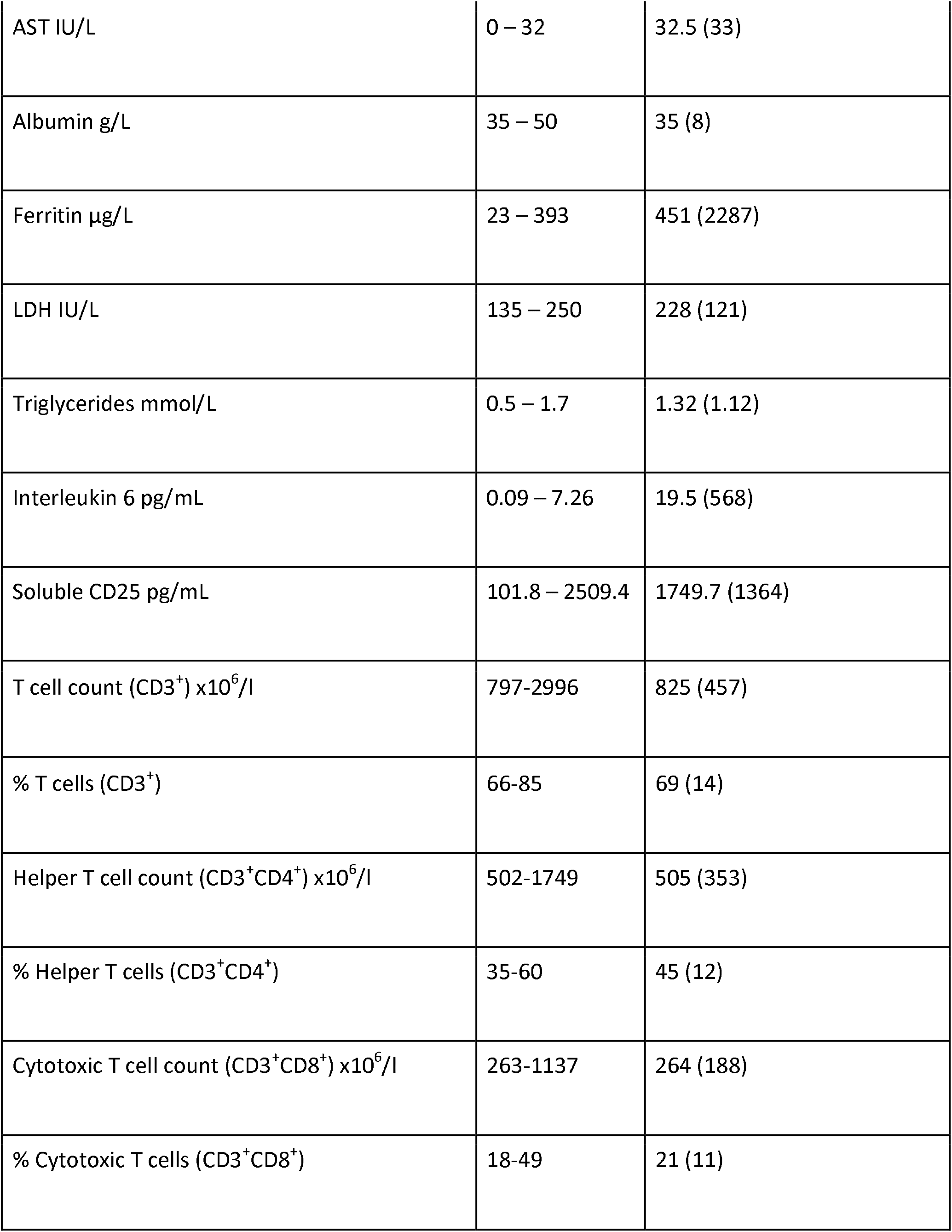
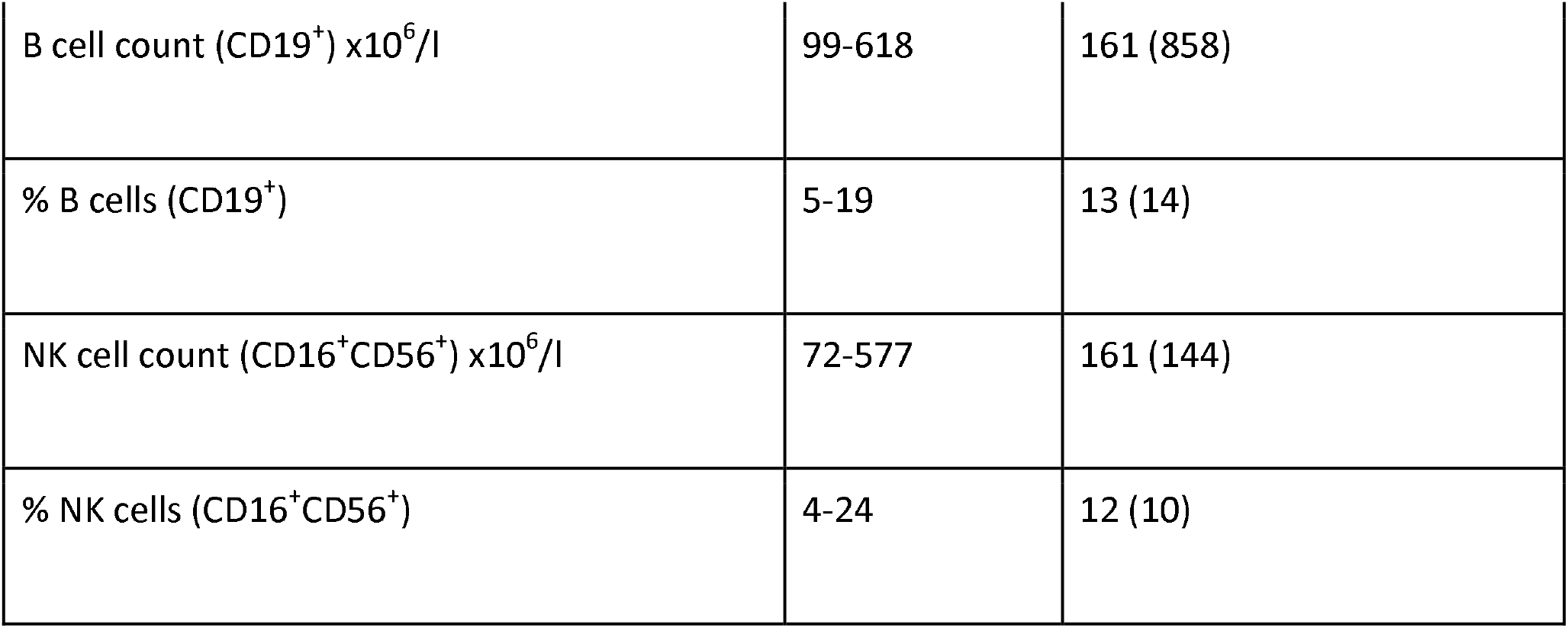
Laboratory Parameters.

### Severe SARS-CoV-2 infection is associated with evidence of profound immune dysregulation

Analysis of routine laboratory tests revealed widespread changes in leukocyte populations with a marked coagulopathy and increased markers of cell turnover and inflammation (**Figure 1**). The degree of lymphopenia significantly increased with disease severity, while marked neutrophilia was seen particularly in those with severe disease. Anaemia with a left shift, characterised by increased red cell distribution width, was also noted with severe disease. Greater disease severity was associated with greater D-dimer and fibrinogen levels, as well as. markers of cellular inflammation (CRP, IL-6, ferritin) and cell turnover (LDH) increased with disease severity.

**Figure 1.**
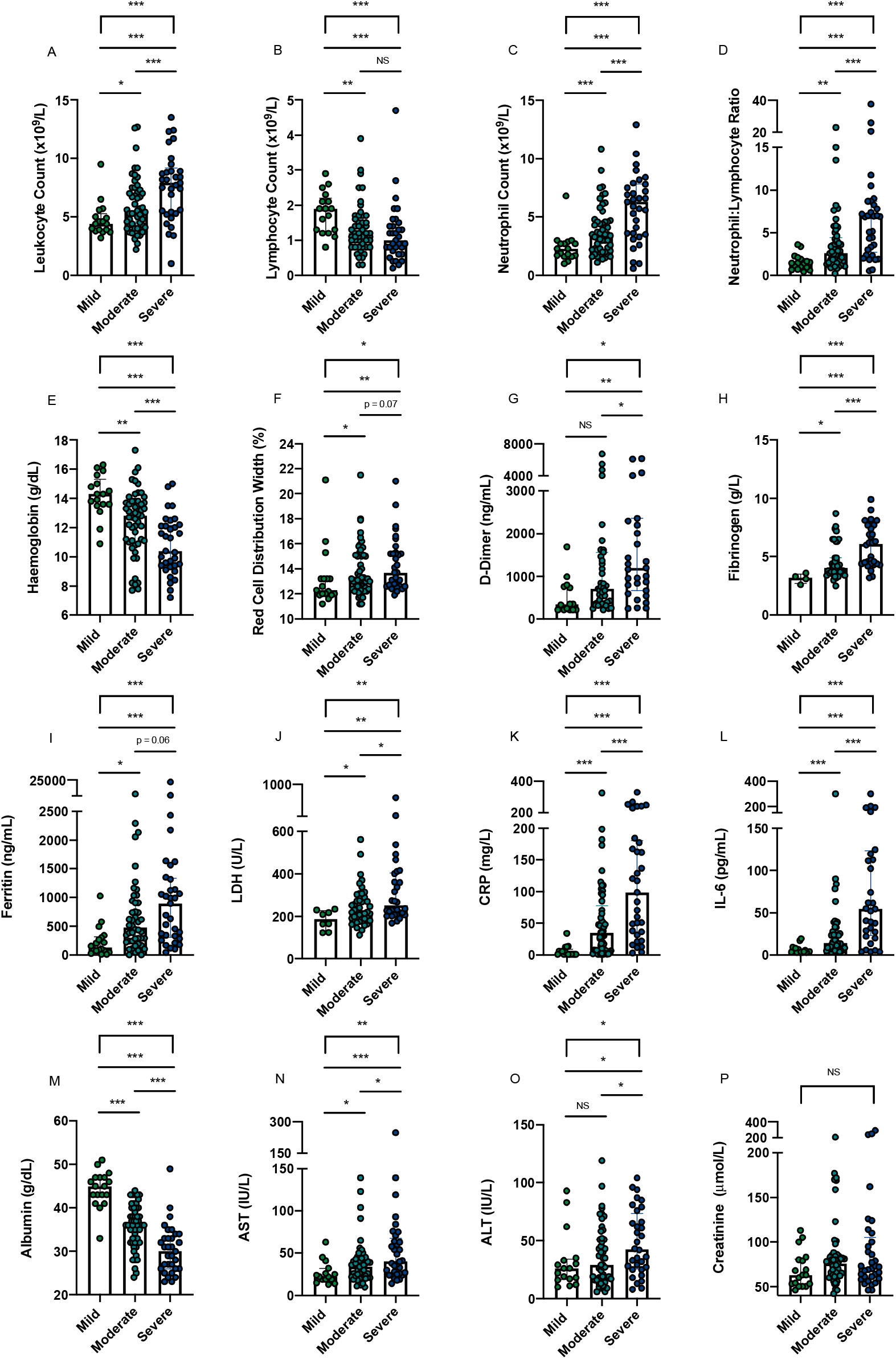
Severity of acute COVID-19 and Markers of Inflammation, Cell Turnover and Coagulation. Severe COVID-19 is accompanied by **(A)** leukocytosis **(B)** lymphopenia **(C)** neutrophil and **(D)** increased neutrophil: lymphocyte ratio in disease (N = 108 total). Severe COVID-19 is also associated with **(E)** lower haemoglobin **(F)** greater red cell distribution width **(G)** increased D-dimer **(H)** increased fibrinogen. Severe COVID-19 was associated with increasing **(I)** LDH **(J)** Ferritin **(K)** CRP **(L)** IL-6, **(M)** lower albumin **(N)** increased AST **(O)** increased ALT. No change in **(P)** creatinine with severity. * p <0.05, ** p <0.01, ***p<0.001, NS = Not Significant

In order to further delineate the panlymphopenia seen on the full blood count, detailed immunophenotyping of lymphoid cells was conducted (**Figure 2**). This confirmed panlymphopenia, with total reduction in total CD3+ cells across all disease severities, most marked in those with severe illness. There was reduction in both CD4+ and CD8+ T cell counts in a similar pattern. On further subset analysis, there was global reduction of naïve CD4+ and CD8+ cells and increased activated CD4+ and CD8+ cells, as well as increased effector CD8+ cells across all disease states. The effect of disease severity was less marked in these subsets, with similar numbers of activated and effector cells across all patients, and similar naïve cell counts in both moderate and severe disease.

**Figure 2.**
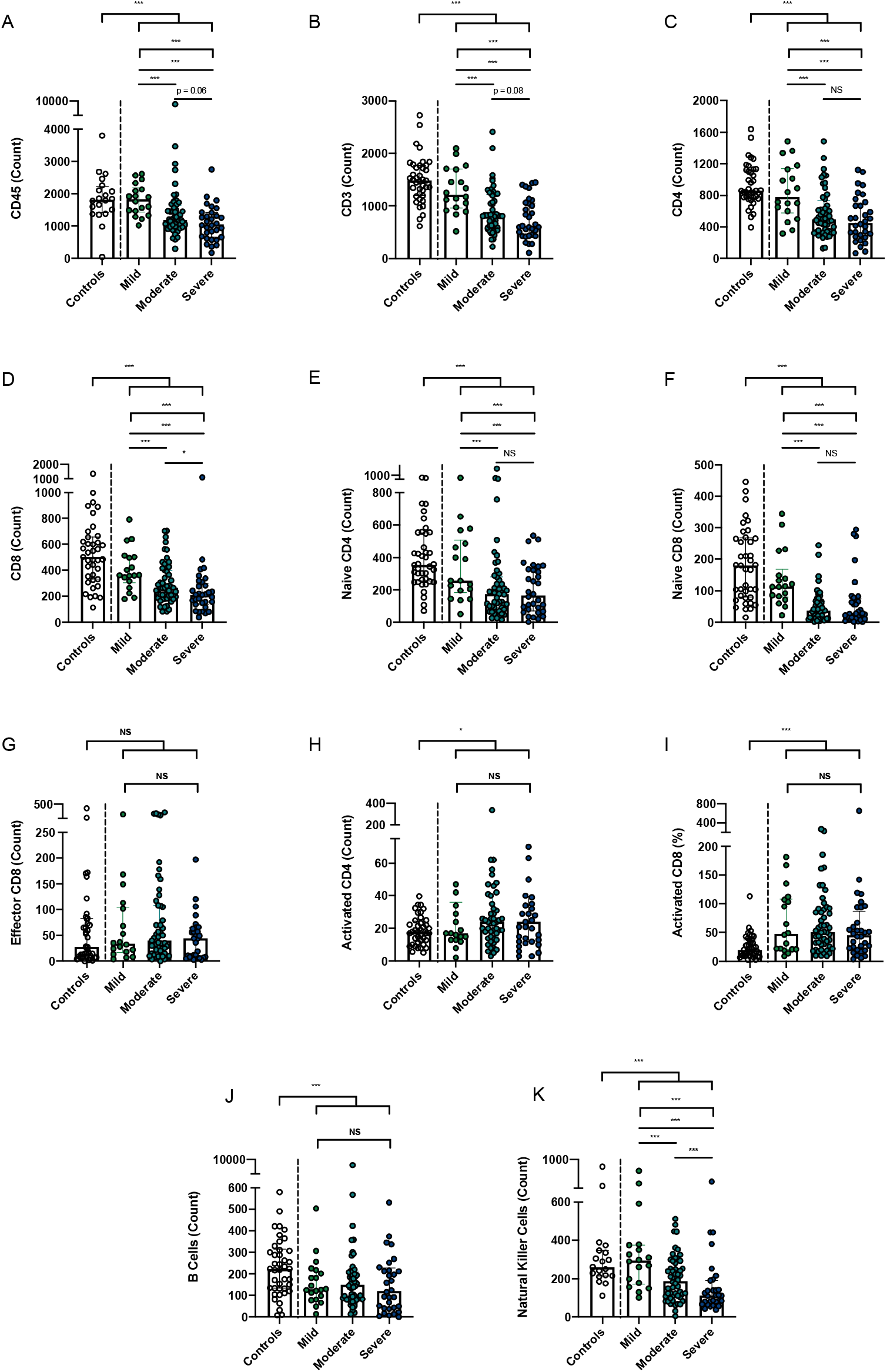
Analysis of Major Lymphoid Subsets Reveals a Widespread Lymphopenia in COVID-19 Disease. Peripheral blood immunophenotyping in those with mild, moderate and severe coronavirus disease revealed significant decreases in **(A)** CD45 positive cells (leukocytes), most pronounced in moderate/severe disease; significant decreases in **(B)** CD3, **(C)** CD4 and **(D)** CD8 cell counts, greatest in those with moderate/severe disease in comparison to controls; **(E)** naïve CD4 and **(F)** CD8 cells both significantly decreased in COVID-19 with increasing disease severity; whilst **(G)** effector CD8 cells were non-significantly elevated, there was a significant expansion in **(H)** activated CD4+ and **(I)** CD8+ T cells, which did not reflect disease severity. Both **(J)** B cells and **(K)** Natural Killer cells were significantly decreased in number in COVID-19. Decreases in Natural Killer cell number reflected disease severity. * p <0.05, ** p <0.01, ***p<0.001, NS = Non-Significant

Detailed immunophenotyping of myeloid populations was also carried out, given the profound neutrophilia noted on initial investigations (**Figure 3**). Marked severity-associated neutrophilia was confirmed. There was loss of CD10 and CD16 expression by neutrophils with increasing disease severity. This finding was confirmed in two ways, looking at both CD10 and CD16 expression on neutrophils as well as the median frequency intensity of both of these markers. In addition to these neutrophil changes, perturbations amongst monocyte populations were found. While absolute number of monocytes was not associated with infection, there was progressive loss of HLA-DR expression by monocytes with increasing disease severity. This was again confirmed by two methods, in a similar manner to neutrophils. There was an increase in proportion of intermediate monocytes in SARS-CoV-2 patients when compared to healthy controls. This increase was most marked in those with mild disease. While there were no significant differences in non-classical monocyte proportions between infected and healthy patients, there were differences across disease severities, with milder patients having a higher proportion when compared to moderate and severe patients. The classical monocyte proportions remained unchanged when compared to healthy controls and when assessed across disease severities.

**Figure 3.**
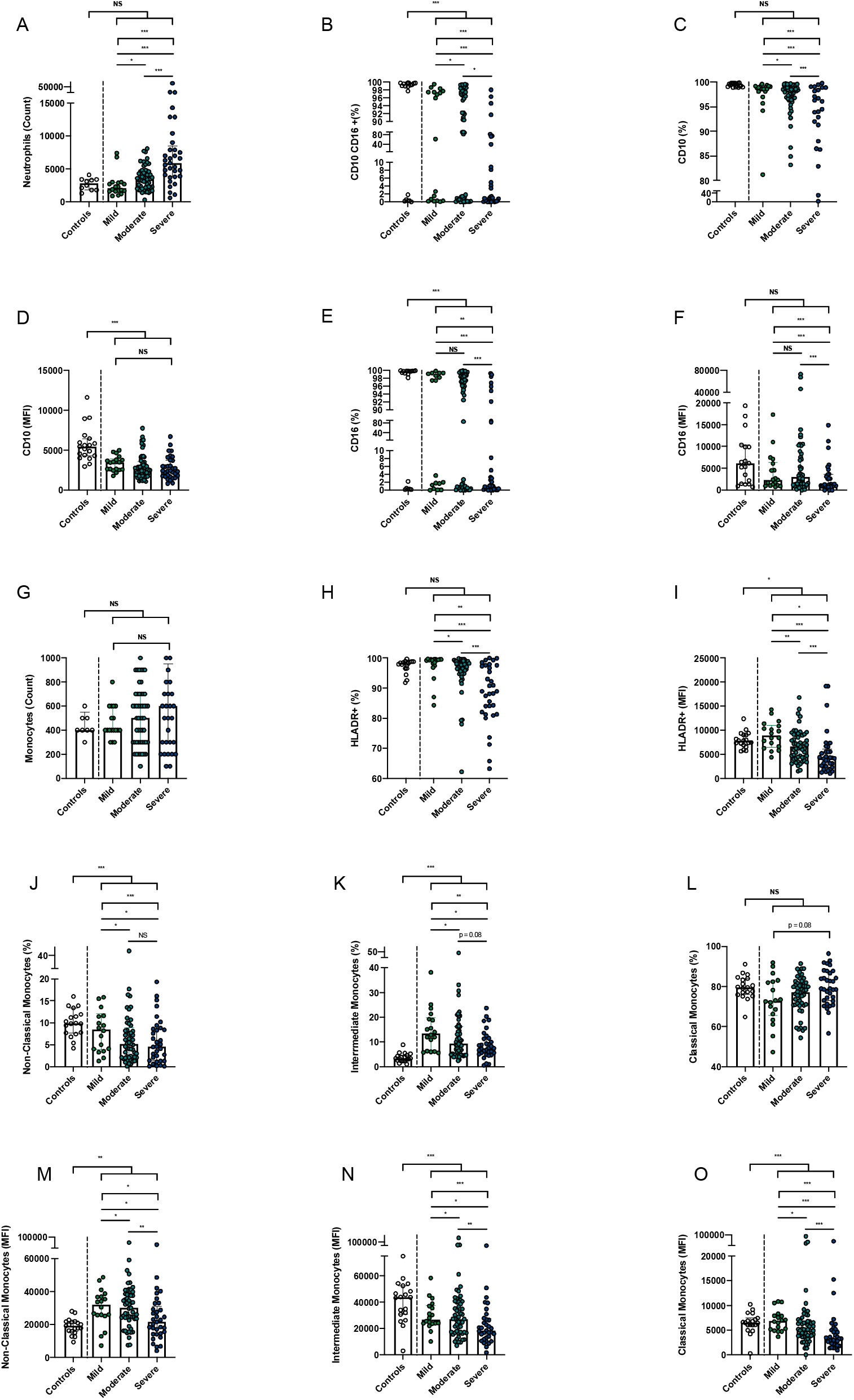
Analysis of Myeloid Cells Reveals a Widespread Immune Dysregulation in COVID-19 Disease. Peripheral blood immunophenotyping in those with mild, moderate and severe coronavirus disease revealed significant changes in neutrophil markers with **(A)** neutrophilia, **(B, E)** reduced neutrophil CD10 expression and **(C)** reduced neutrophil CD16 expression as well **(D, F)** reduced Mean Fluorescence Index (MFI) of these markers, most pronounced in those with severe COVID-19 disease. **(G)** Monocyte numbers were not significantly altered, but there were significant changes in monocyte subsets, with **(H)** reduced HLA-DR+ monocytes, **(J)** lower number of non-classical monocytes, **(K)** increased intermediate monocytes **(L)** no change in classical monocytes. The MFI changes are shown in **(M-O)**. Monocyte subpopulations are shown as proportions of total monocyte population, while neutrophil CD10+ and CD16+ populations are shown as a proportion of total neutrophil population, based on flow cytometry gating. * p <0.05, ** p <0.01, ***p<0.001, NS = Non-Significant

We then proceeded to analyse between-group differences in those with severe COVID-19 in comparison to those with mild or moderate disease. Univariate analysis of the 71 variables in our clinical and laboratory dataset identified a series of 15 factors associated with severe COVID-19 that remained statistically significant when corrected for false discovery rate (**Figure 4A**). These are ranked by strength of association, with strong associations seen with some of the differences noted in our earlier analysis. Specifically, elevation of the acute phase reactants CRP and IL-6 were closely associated with severe disease, with increased soluble CD25 also seen with severe disease. Furthermore, reduced expression of the neutrophil marker CD10 and monocyte marker HLA-DR were strongly associated with severe disease. No lymphocyte marker approached significance. Chi-squared testing of categorical variables ranked homelessness, male gender, higher clinical frailty scores and being a current smoker as the categorical variables most strongly associated with severe COVID-19, although no association met the threshold for statistically significance (**Figure 4B**).

**Figure 4:**
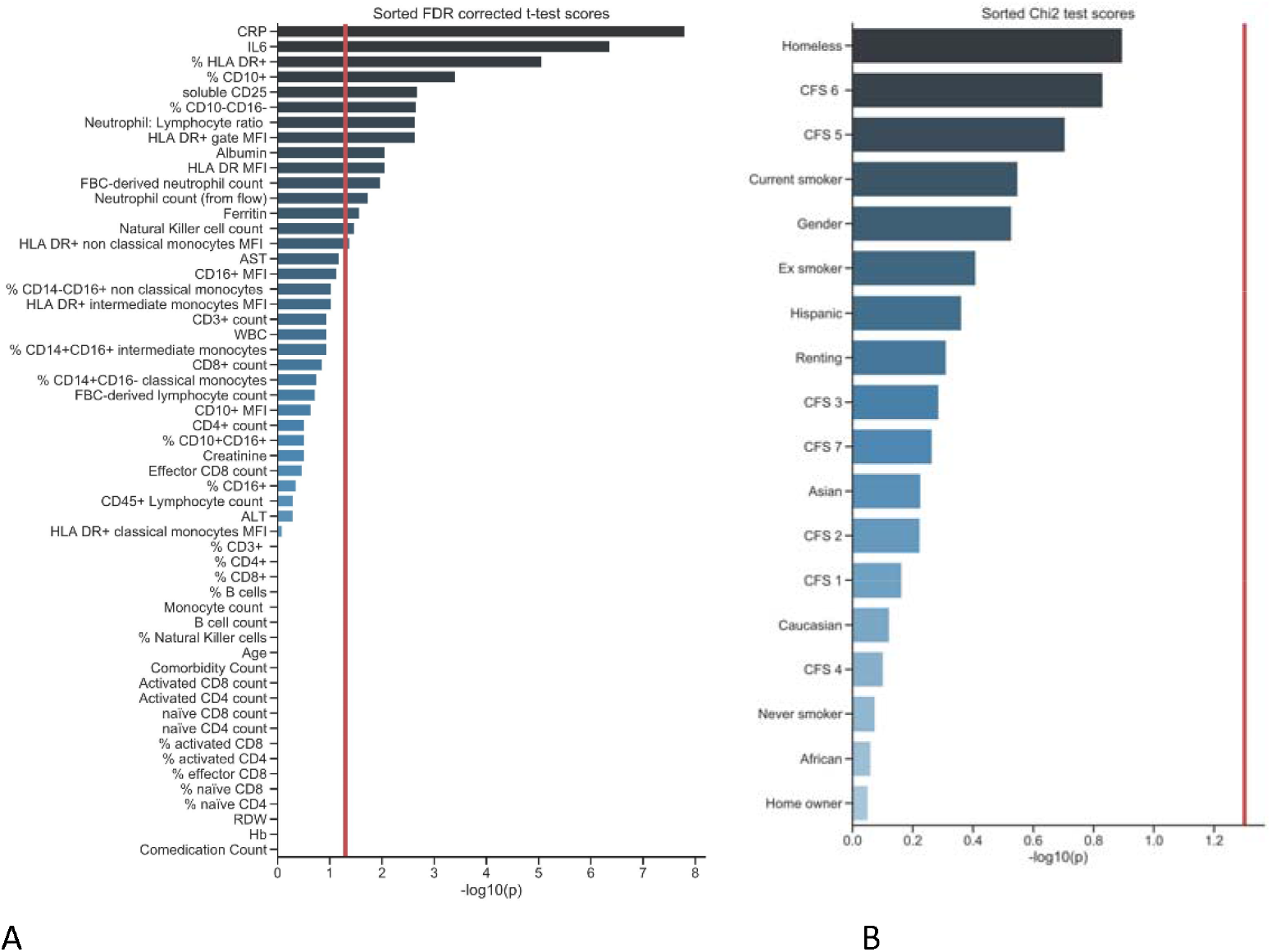
Univariate analysis of numerical demographic, clinical and immunological variables associated with severe COVID-19. **(A)**. t-test scores for differences in means between patients with severe and non-severe COVID-19 are indicated in the bar graph, corrected for false discovery rate using Benjamini–Yekutieli procedure The scores were derived from p-values by calculating -log10 (p-value). The red line indicates threshold of statistically significance (p<0.05). Univariate analysis of categorical variables associated with severe COVID-19 **(B)** Sorted Chi-squared test scores were used to test for significant associations between categorical variables and severe COVID-19. The red line indicates threshold of statistically significance (p<0.05).

### Machine learning identifies and ranks predictors of severity

Having identified strong associations between specific individual immunological features and severity, we utilised multivariate analysis to establish the relative importance of the clinical and immunological parameters when adjusted for high-dimensional interactions. To establish this, we fitted a logistic regression model with elastic net penalty to the data. Our model demonstrated a high sensitivity and specificity of prediction for severity. The area under the receiving operating characteristic (AUROC) curve was 0.93 on both training and test sets, indicating excellent model predictive performance and absence of overfitting (**Figure 5B**). These results were confirmed with a permutation test resulting in statistically significant output (**Supplemental Figure 3A**; p<0.01). A precision-recall curve was also applied to estimate the model’s predictive discriminating performance due to the class imbalance in the ICU outcome measure (39 versus 69 cases). The average precision (AP) score was 0.89 on the training set and 0.88 on the test set (**Figure 5B**). Both of these scores indicate excellent generalisation properties for this model. The model’s true positive and true negative rates were assessed on training and test sets, visualized by confusion matrices (**Supplemental Figure 3B**), which indicate the model’s excellent discrimination between classes and absence of overfitting.

**Figure 5:**
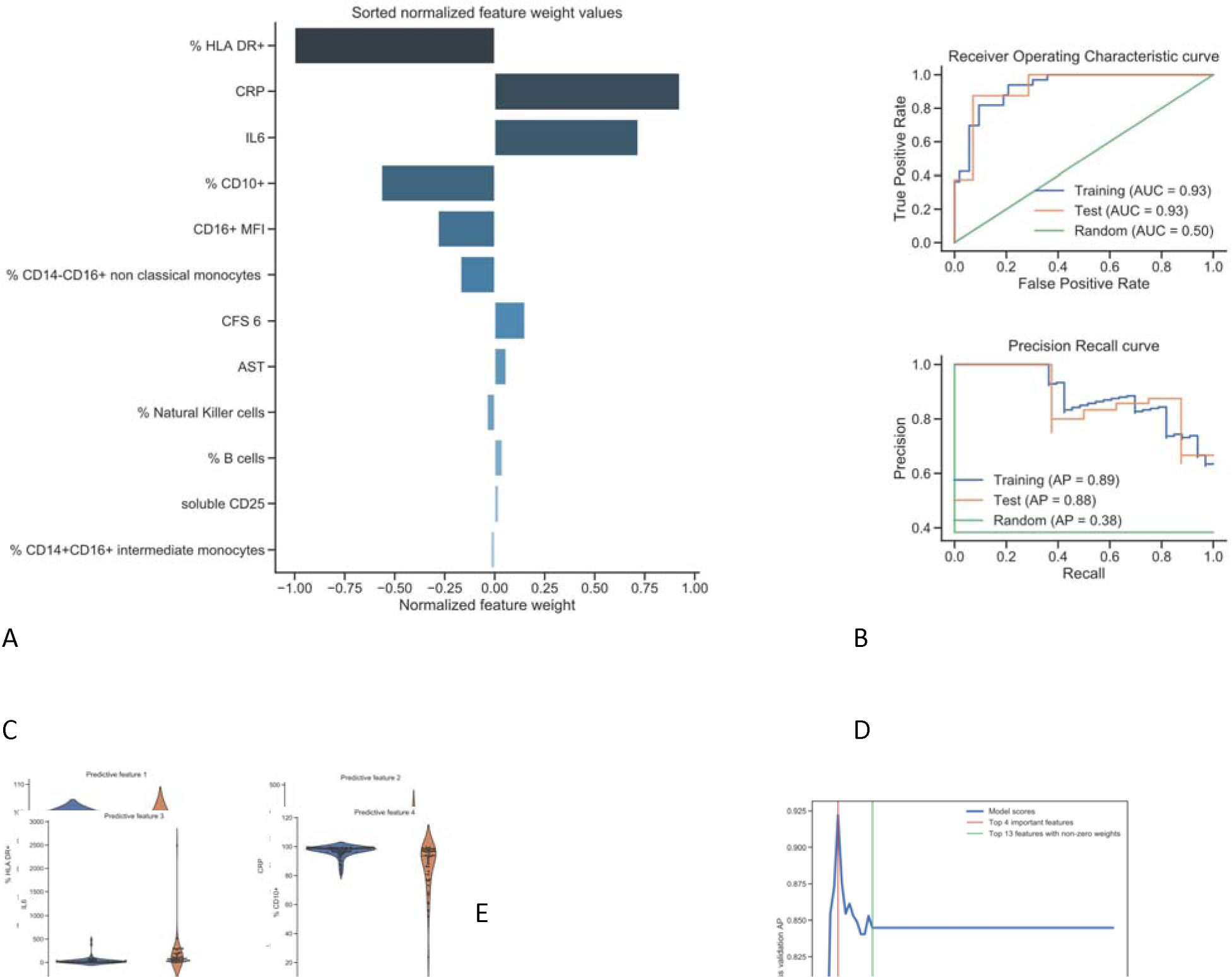
Machine learning modelling of variables identifies immunological features as the strongest predictors of severe COVID-19. **(A)** Sorted normalised feature weight values of variables identified by the model as predictive of progression to severe COVID-19 **(B)** Model performance for training and test sets assessed using receiver operating characteristic (ROC) curves and precision-recall curves. **(C)** Violin plots representing the difference of values between severe and non-severe COVID for the four features with the highest weight values in the predictive model **(D)** Predictive power of top four variables **(E)** Correlation matric of predictive variables within the model, analysed using Spearman correlation Blue indicates negative correlation between variables, red indicates positive correlation. Darker colour indicates a stronger association. COVID-19. Data was fitted to a logistic regression model with elastic net penalty. For the training set, the hyperparameters were optimized using a 10-fold cross-validation procedure and an exhaustive grid search on a training subset comprising 80% of the data. The remaining 20% of the data was used for the test set.

When applied to our integrated dataset, the machine learning multivariate approach identified 12 variables *predictive* of progression to severe COVID-19 (i.e. with associated weights of non-zero values) when considered in interaction with all 71 variables. Normalised weight values associated with the top features of the model are shown in **Figure 5A**. The top four features stand out as most important with normalised weight values between 1.0 and 0.56. (**Figure 5C**)

The normalised weight values of subsequent features were lower (0.28 to 0.01). The top feature predictive of severe COVID-19 was a reduced proportion of HLA-DR^+^ monocytes. The next most important features were higher CRP and IL-6 levels, and a lower percentage of CD10^+^ neutrophils (**Figure 5A**). Strikingly, a combination of these four features alone could predict COVID-19 severity with an average precision score of 0.925. Additional immunological features associated with severe COVID-19 with lower weight values included lower CD16 expression by neutrophils, lower percentages of non-classical (CD14^-^CD16^+^) and intermediate (CD14^+^CD16^+^) monocytes and lower numbers of NK cells (**Figure 5A and Supplemental Figure 4A, 4B, 4H**). Other parameters associated with severe COVID^-^19 were a high clinical frailty score of 6, higher AST levels, higher B cell counts and higher sCD25 levels (**Figure 5A and Supplemental Figures 4C-G**). Importantly, introducing additional variables to the four highest-ranked features did not improve the predictive ability of the model (**Figure 5D**).

Potential relationships between each variable identified in the machine learning analysis were also investigated using a clustered Spearman correlation matrix (**Figure 5E**). Unsurprisingly, there was a positive correlation between levels of CRP and IL-6, with both of these markers negatively correlated with levels of HLA-DR^+^ monocytes and CD10^+^ neutrophils. Levels of HLA-DR^+^ monocytes and CD10^+^ neutrophils correlated positively with each other. There was a positive correlation between IL-6 and CRP with AST and sCD25. Proportional changes in non-classical and intermediate monocytes were also strongly associated with each other.

## Discussion

COVID-19 has presented an enormous challenge to human health and society. The dramatic variability in clinical outcomes of SARS-CoV-2 infections has been proposed to be driven by variation in immune responses.^10^ Known risk factors for severe COVID-19 include older age, male gender, high BMI, and co-morbidities such as Type 2 diabetes and cardiovascular disease.^28^ The nature of the early stages of immune dysregulation in COVID-19, and how predictive it may be of disease severity in relation to these other important risk factors, is an area that still warrants further exploration.

The data presented here on 108 patients with SARS-CoV-2 infection confirms experimental and research laboratory work on the characteristics of severe infection. Notably, it includes patients across the spectrum of disease severities; 69 (60%) of patients required supplemental oxygen therapy, 39 (36%) progressing to severe COVID-19, and 17 (16%) had mild disease managed in ambulatory care. In addition, and importantly, our methodology is suitable for use in a diagnostic immunology laboratory. Our cohort is broadly reflective of those reported elsewhere, with a median age of 61, more males than females requiring admission and a mortality rate of 9% which was higher for men.^24^ The similarities to cohorts reported elsewhere are crucial, given that early attempts to describe the immunopathological signatures of COVID-19 have been poorly applicable across populations.^25-27^ We used a whole blood flow protocol, with minimal handling and analysis within 4 hours of blood draw. This minimized artefactual alterations in myeloid populations. Patients presented to hospital and had their bloods drawn a median of 2 days prior to peak COVID-19 as defined by peak oxygen requirement, giving us insight into the mechanisms leading to severe respiratory distress.

Severe disease in our cohort is characterised by markedly elevated inflammatory markers and a shift towards emergency myelopoiesis. Those with severe illness demonstrate increased inflammatory cytokines IL-6, CRP and soluble CD25. This works builds on several studies that have highlighted the importance of elevated CRP, IL-6 and the acute phase reactants in COVID-19.^29^ We also see a strong association with immature myeloid cells, both neutrophils and monocytes, with severe disease. This shift in myelopoiesis has been highlighted previously^30^, with left-shift of myeloid cells along with increased cytokine production has been seen in early infection.^31^ We also demonstrate the emergence of anaemia and increased RDW in severe disease, suggestive of emergency haematopoiesis. Indeed, increased RDW over the course of inpatient stay has been associated with increased COVID-19 mortality.^32^

Our data demonstrates evidence of inflammation and immune perturbation early in progression to severe COVID-19, with the emergence of a quartet of markers strongly associated with progression to severe disease. These markers are seen before the onset of peak clinical illness in our cohort. The strongest single predictor of severity on multivariate analysis was a reduced proportion of HLA-DR^+^ monocytes. Reductions in HLA-DR^+^ monocytes and reduced HLA-DR expression are appear to be an early predictor of poor outcome across many scenarios other than COVID-19.^33-36^ Increasing data have emerged showing reduced monocyte HLA-DR expression is associated with critical illness in COVID-19.^16, 20, 37^ The second member of the prediction quartet identified by this study is a proportional reduction in CD10^+^ neutrophils, another marker of emergency myelopoiesis. CD10 is a cell membrane metalloprotein that serves as a marker of neutrophil maturity.^38^ Reduced neutrophil CD10 expression is suggestive of an immature and proinflammatory population. Immature neutrophils, characterised by lower or absent CD10, have been reported to be immunostimulatory by promoting T-cell proliferation, survival and IFNγ production and have been associated with poor prognosis in sepsis.^39, 40^ Immature neutrophils characterised by low CD10+ have recently been associated with aggressive SARS-CoV-2 infection.^20^

The final two members of the predictive quartet are the acute phase reactants IL-6 and CRP. Elevated levels of IL-6 (and downstream protein CRP) are well-described as predictors of poor outcome in COVID-19.^41-44^ Elegant studies have highlighted a potent pro-inflammatory cytokine response, with prominent IL-6 elaboration, accompanied by a curious blunted type I and III interferon responses that seems unique to SARS-CoV-2 infection.^28, 45^ IL-6 also appears to be an important driver of monocyte HLA-DR loss, placing this cytokine at the centre of the immunopathological signature of COVID-19.^16^

Surprisingly, only a single categorical clinical variable, higher clinical frailty scores, emerged on multivariate analyses as associated with severity. The clinical frailty score captures biological, rather than chronological age and is a predictor of mortality in a wide-range of conditions.^47, 48^ Remarkably, a large number of other expected variables, including age and gender, failed to emerge as risk factors for severity in our cohort – however, this must be qualified by the fact that our cohort included few individuals with mild disease and does not rule out a role for these factors as risk factors for hospitalisation, for example.

This is a single centre study in a largely Caucasian Irish cohort of predominantly hospitalised patients. Validation of these findings in other centres and with larger samples earlier in the course of disease is desirable.

Understanding of the mechanisms causing some individuals with SARS-COV-2 infection to progress to severe disease is required to inform therapeutic design. Through robust analysis of extensive demographic, clinical and immunological parameters captured early in disease trajectory, we rank these parameters and adjust them for high-dimensional interactions and report a signature predictive of severity which is characterised by inflammation (elevated IL-6 and CRP) and emergency myelopoiesis (reduced neutrophil maturity and monocyte HLA-DR expression).

This study builds on prior work describing severe COVID-19 infection, demonstrating that these features of emergency myelopoiesis develop prior to the point of peak illness and are of central importance in predicting progression to severe COVID-19. We include patients with mild disease who did not require hospitalisation; these patients represent a heretofore understudied cohort. Furthermore, we confirm that these approaches can be delivered in an accredited clinical laboratory and have the potential to be developed into routine tests. This is of particular importance given the need for early identication of individuals who may benefit from therapeutic interventions.^22^ Our work emphasises the central role of expanded immature neutrophil and monocyte populations in the setting of COVID-19 which may be similar to the expanded low density granulocyte and monocytic myeloid-derived suppressive cells seen in sepsis.^46^

## Data Availability

All relevant data is included in the manuscript. Further queries can be addressed to the corresponding author.

## Acknowledgements

The authors would like to thank all patients who took part in this study, our colleagues who provided care for patients and St James’s Hospital and the Wellcome CRF who supported the study. L.T. has been awarded the Irish Clinical Academic Training (ICAT) Programme, supported by the Wellcome Trust and the Health Research Board (Grant Number 203930/B/16/Z), the Health Service Executive, National Doctors Training and Planning and the Health and Social Care, Research and Development Division, Northern Ireland. R.M.M. has been awarded a Science Foundation Ireland (SFI) Investigator Award (15/IA/3041). N.C. and C.N.C. are part-funded by an SFI grant, Grant Code 20/SPP/3685. The funders had no role in study design, data collection and analysis, decision to publish, or preparation of the manuscript.

## Author Contributions

Conceptualization: L.T., J.D., N.M.B., C.O’F., N.C. and C.N.C.; Methodology: L.T., A.H.D., S.I. and R.M.M.; Software: S.I. and A.W.; Validation: A.N., S.I., J.D., R.K., D.H., A.M., D.L., J.S., A.W. and R.M.M.; Formal Analysis: L.T., A.H.D., S.I. and A.W.; Investigation: L.T., A.H.D., A.N., J.D., R.K., D.H., A.M., D.L., K.R., J.S., M.A. and J.H.K.; Resources: M.H., D.B., P.B., C.L.B., C.D., R.O’R., A.M.M., C.Ba., I.M.L. and C.Be.; Data Curation: L.T., A.H.D., A.N., S.I., J.D. and A.W.; Writing-Original Draft: L.T., A.H.D., A.N., N.M.B., C.O’F., N.C. and C.N.C; Writing-Review & Editing: D.B., P.B., C.L.B., C.D., R.O’R., A.M.M., C.Ba., I.M.L. and C.Be.; Visualization: A.N., S.I., J.S., A.W., N.M.B., N.C. and C.N.C.; Supervision: J.D., A.W., R.M.M., C.Be., N.M.B., C.O’F, N.C. and C.N.C.; Funding Acquisition: L.T., R.M.M. and N.C.

## STAR Methods

| REAGENT or RESOURCE | SOURCE | IDENTIFIER |
| --- | --- | --- |
| **Antibodies (Fluorophore, clone)** |  |  |
| CD45 (V500-C, 2D1) | BD Biosciences | Cat#655873 |
| CD8 (V450, RPA-T8) | BD Biosciences | Cat#560347 |
| CD3 (APC-H7, SK7) | BD Biosciences | Cat#641415 |
| CD4 (PerCP/CY5.5, SK3) | BD Biosciences | Cat#332772 |
| CD45RA (PE, -) | BD Biosciences | Cat#556627 |
| CD27 (FITC, -) | BD Biosciences | Cat#555440 |
| CD197 (Alexa Fluor 657, 150503) | BD Biosciences | Cat#560816 |
| HLA DR (FITC, L243) | BD Biosciences | Cat#347400 |
| CD38 (APC, HB-7) | BD Biosciences | Cat#345807 |
| CD14 (APC, MΦP9 ) | BD Biosciences | Cat#345787 |
| CD16 (PE, B73.1) | BD Biosciences | Cat#332779 |
| CD10 (APC, HI10a) | BD Biosciences | Cat#332777 |
| **Commercial Assays** |  |  |
| IL-6 ELISA | R&D Systems | Cat#D6050 |
| sCD25 ELISA | R&D Systems | Cat#DR2A00 |
| BD TruCount | BD Biosciences | Cat#340334 |
| **Software and Algorithms** |  |  |
| Python 3.7 | Python | www.python.org |
| BD FACSDiva v8 | BD BioSciences |  |
| FLO Jo v10 | BD | www.flojo.com |

### Subject Details

This study was carried out in accordance with the Code of Ethics of the World Medical Association.^49^ Ethical approval was granted by the Tallaght University Hospital / St. James’s Hospital Joint Research Ethics Committee (reference REC 2020-03). Informed consent was obtained from all participants. We recruited patients with a positive real-time PCR test for SARS-CoV-2 on nasopharyngeal aspirates at our institution over a two-month period (**Supplemental Figure 1**). 71 demographic, clinical and immunological measures were recorded for participants enrolled in this study (**Supplemental Table 1**). These included social and medical history (smoking status, alcohol and drug use, housing status, co-morbidities, medication use and self-reported date of symptom onset), and clinical frailty scores (adjudicated by the admitting physician).^50^ Routine haematological and biochemical laboratory results at the earliest time point after the first positive SARS-CoV-2 test, radiologist-reported chest X-ray results, and clinical parameters including peak oxygen requirement and peak CRP value (within fourteen days of positive SARS-CoV-2 test), assessment for ICU admission, admission to ICU and outcome were extracted from the electronic patient record. Mild COVID-19 was defined as no requirement for hospital admission (WHO scale 1 and 2), moderate COVID-19 as requirement for hospital admission (WHO scale 3 and 4) and severe COVID-19 as requirement for high-level respiratory support (high flow oxygen, invasive ventilation (WHO scale 5 and 6).

### Methods Details

Immunophenotyping was carried out on fresh whole EDTA treated blood and samples were analysed on a FACS Canto II Flow Cytometer (BD San Jose USA), using BD DIVA v8 and FLO Jo v10 software. T cell (CD4^+^ and CD8^+^), B cell (CD19^+^) and Natural Killer cell (CD16^+^CD56^+^) percentages and absolute counts were generated by standard methods (BD TruCount). Naive (CD27^+^) and Effector (CD27-) T cells were characterised using CD27, CD45RA and CD197 antibodies. T cell activation was assessed by CD38 and HLA-DR expression. These assays were previously validated and accredited in line with ISO15189 standards. Classical, intermediate and non-classical monocytes were characterised by CD14 and CD16 expression. Proportions of these subpopulations were reported as percentages of the total monocyte population. Neutrophil functional maturity was interrogated by evaluating CD10 and CD16 expression. Proportions of these populations were reported as percentages of the total neutrophil population. IL-6 and soluble CD25 (sCD25) levels were measured in serum by ELISA (R&D Systems, Minneapolis, USA). Results were compared to a cohort of 40 pre-pandemic healthy controls, 20 of whom had extensive analysis performed for lymphoid and myeloid cell subsets, with 20 controls having extensive lymphoid analysis only.

### Statistical Analysis

Between group differences were examined in the first instance using standard univariate statistics (t-tests, Wilcoxon rank-sum and chi-square tests as appropriate). In order to compare differences across the three groups of increasing COVID-19 severity, we used Kruscal-Wallis testing with a Dunn’s post-hoc test to estimate p-values. An alpha level of p<0.05 was considered statistically significant.

We analysed a total of 71 demographic, clinical, laboratory and immunophenotyping parameters (independent variables) to analyse factors associated with severe COVID-19 (**Supplemental Table 1**). All data analysis, visualizations and modelling were performed using open source packages available in Python 3.7.^51, 52^ For univariate analysis, a FDR corrected t-test was applied on power transformed numerical variables. The power transformation was implemented using Yeo-Johnson transformation to ensure normal distribution of variables. FDR-correction was implemented using Benjamini-Yekuteli procedure.^53^ Categorical variables were tested using Chi^2^-test. An elastic net regularized logistic regression model was used for multivariate analysis and implemented for simultaneous feature selection and ICU status prediction.^54^ To train each model, the hyperparameters were optimized using a 10-fold cross-validation procedure and an exhaustive grid search on a training subset comprising 80% of the data. Generalization of the trained model was then assessed using the remaining 20% of the data as a test set.

## Supplemental material titles and legends

**Supplemental Table 1:**
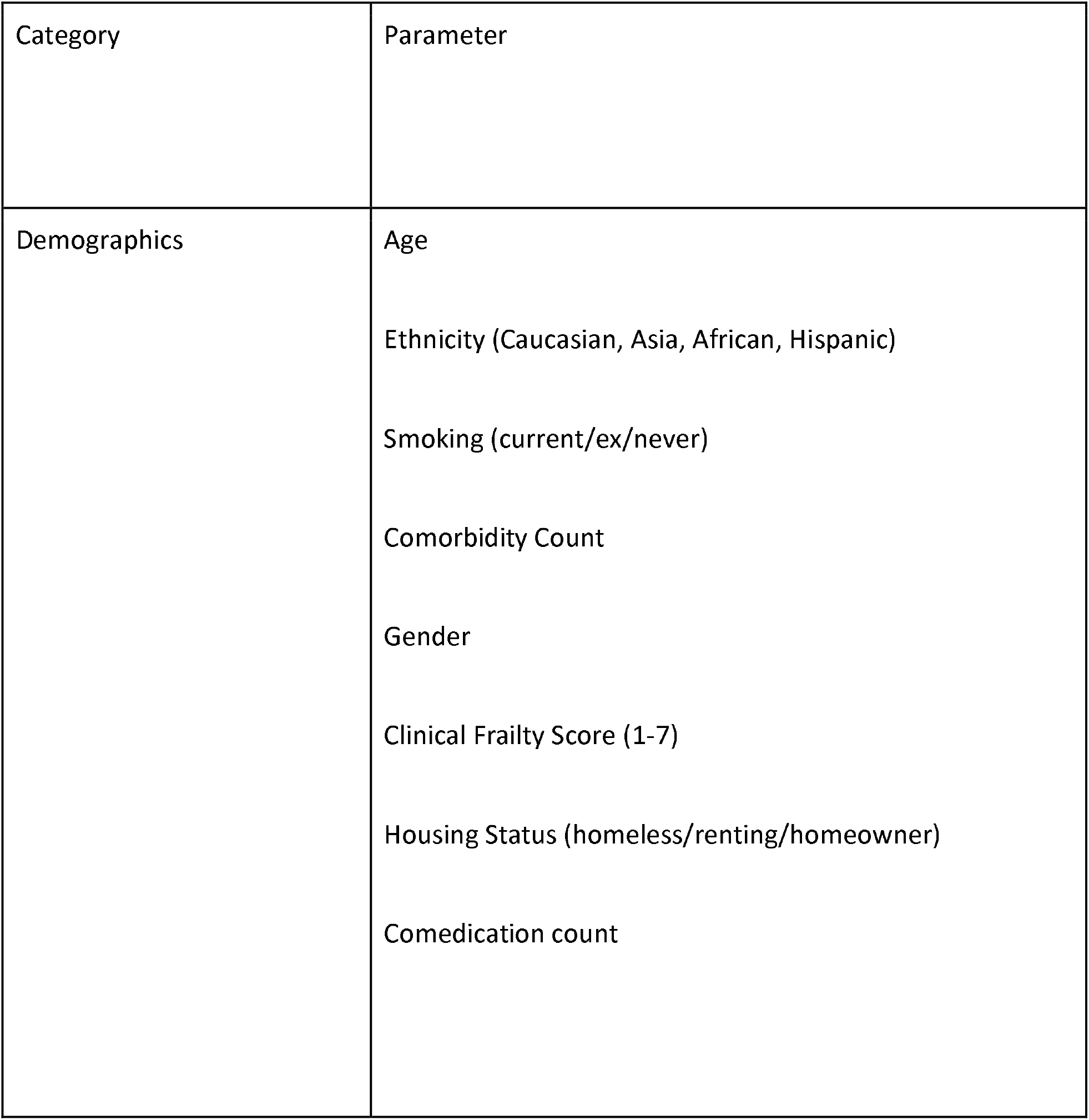
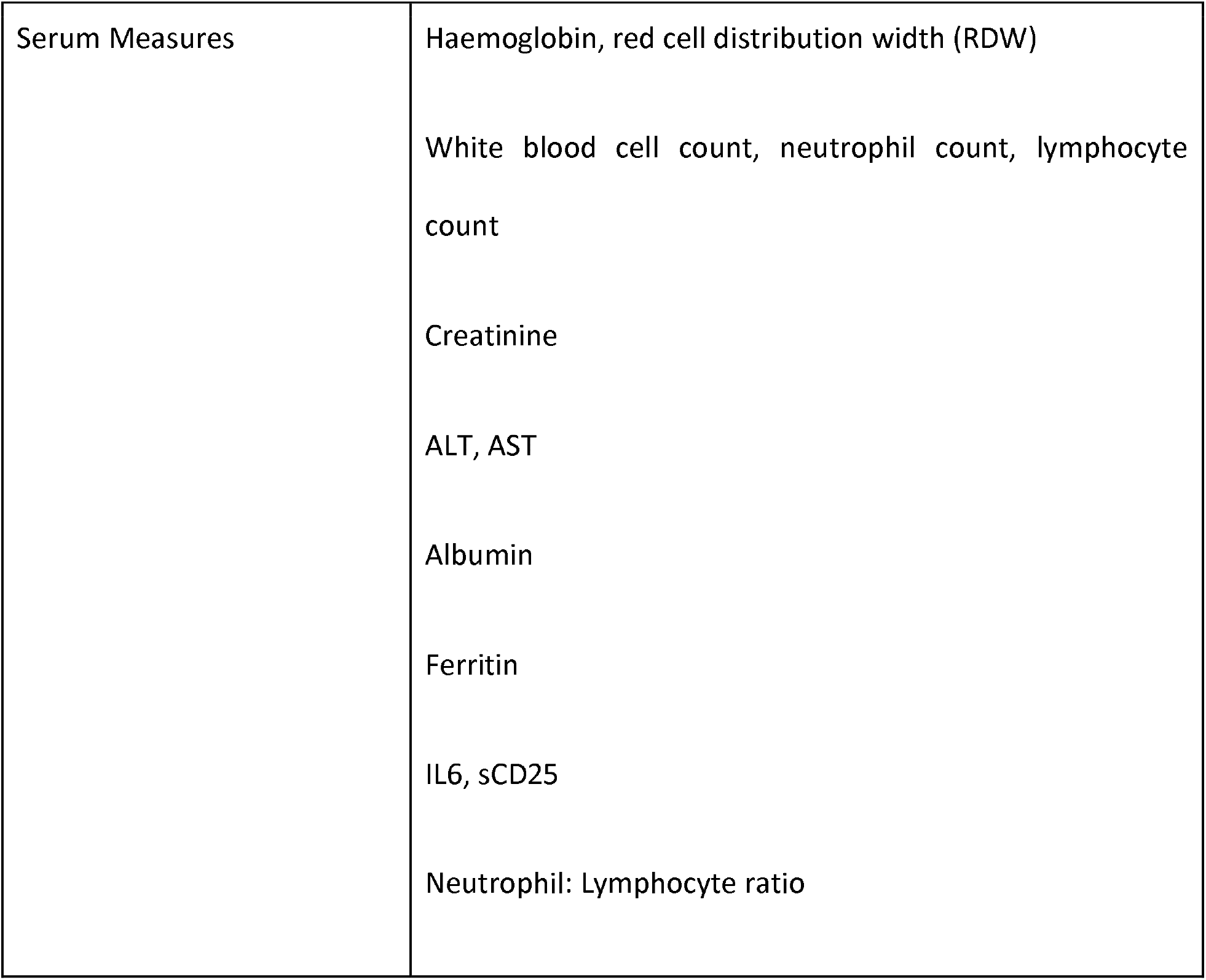
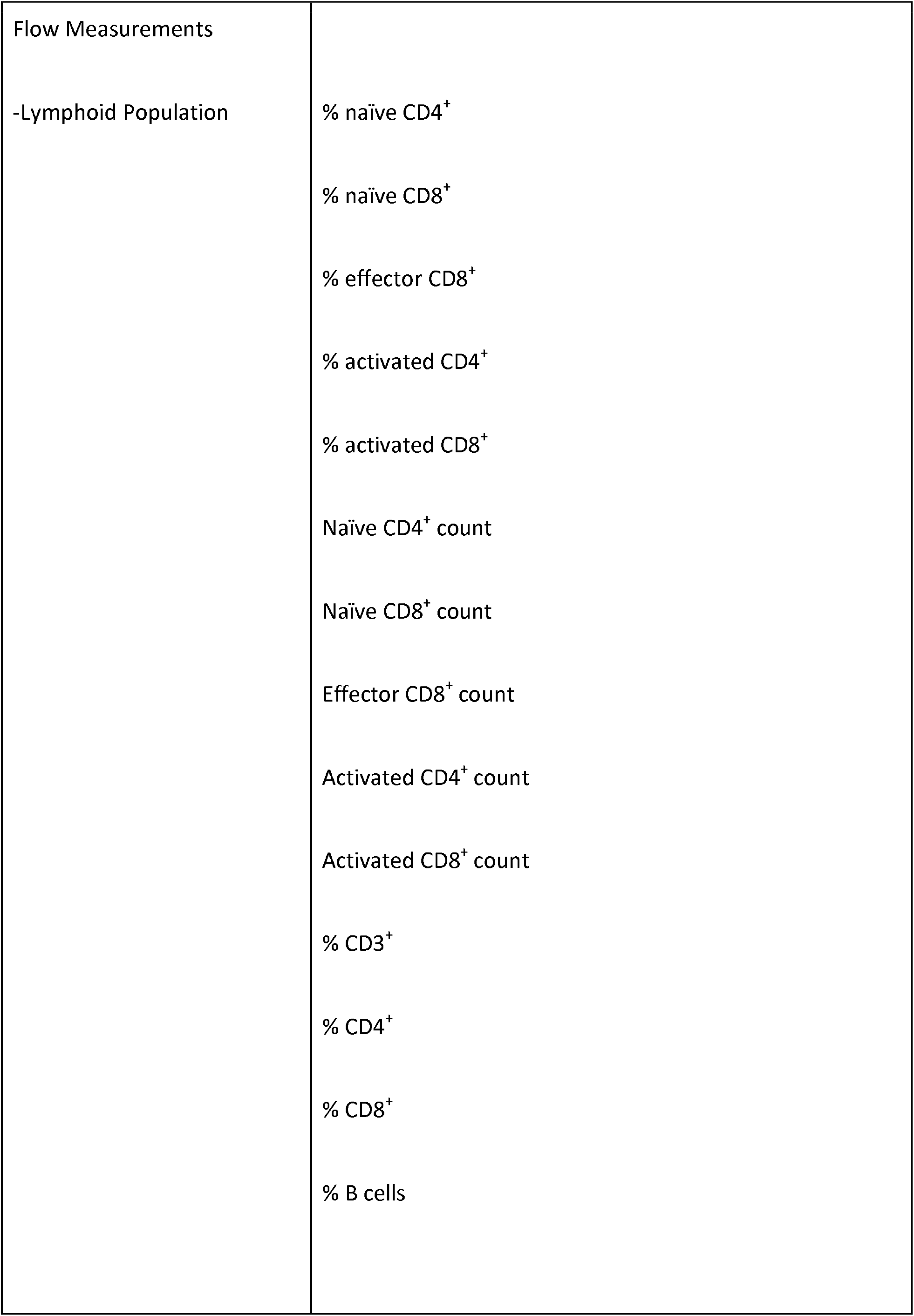
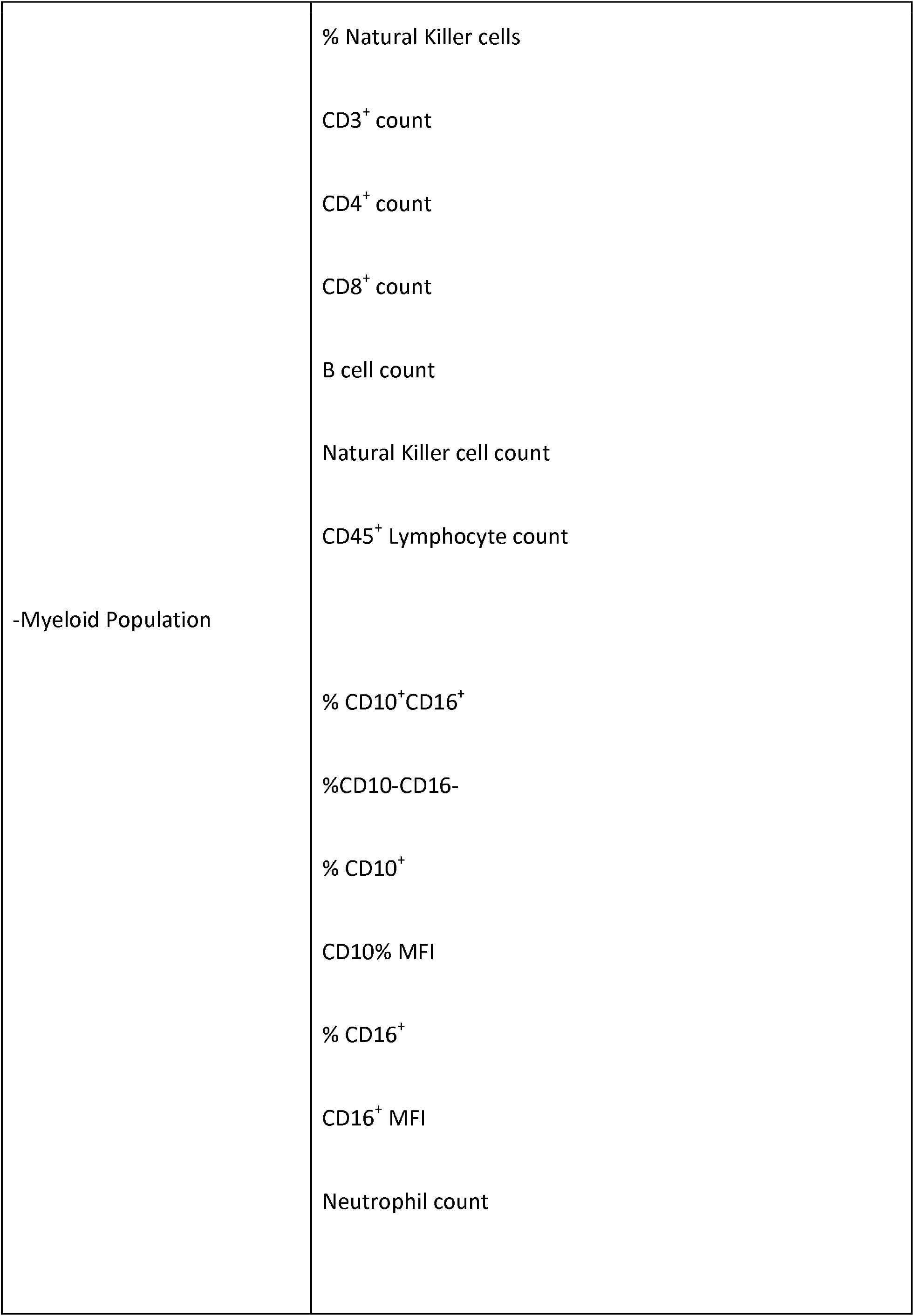
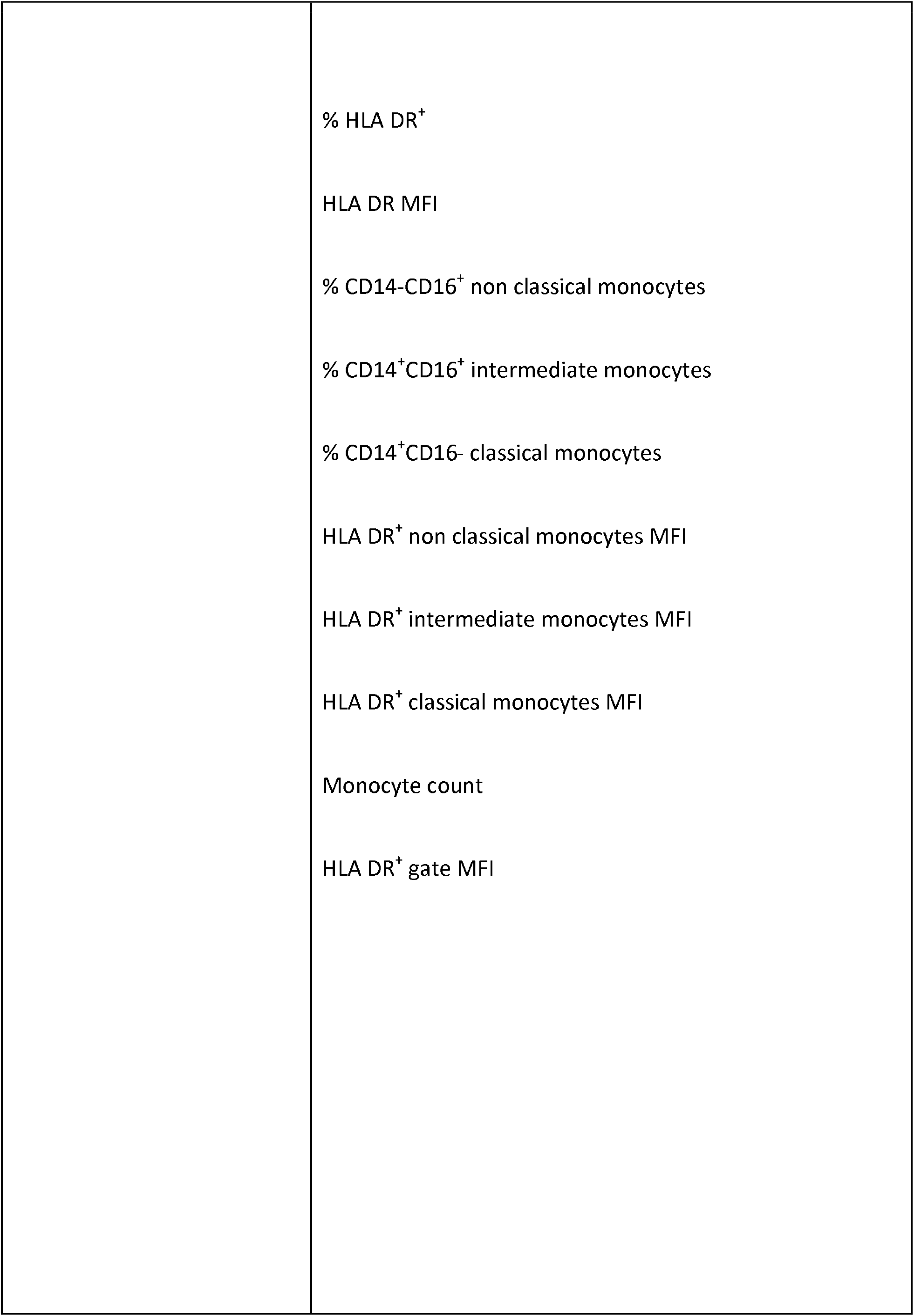
71 parameters included in analysis.

**Supplemental Table 2:** Timing of samples.

| Time period | Median days (range) |
| --- | --- |
| Symptom onset to first blood draw | 6 (1 – 15) |
| First blood draw to peak oxygen requirement (n 69) | 2 (0 – 14) |
| Blood draw for immunophenotyping to peak oxygen requirement (n 69) | 1 (13 pre to 33 post) |

**Supplemental Figure 1: Patient enrolment flow chart**

**Supplemental Figure 2: Flow cytometry gating strategy. (A)** Naïve and effector T lymphocytes **(B)** activated T lymphocytes **(C)** neutrophils **(D)** monocytes

**Supplemental Figure 3: Performance of machine learning model. (A)** Permutation test of model **(B)** Confusion matrices, showing performance of model on training and test sets

**Supplemental Figure 4: Violin plots of predictive features 5 – 12. (A)** CD16 MFI **(B)** Proportion of non-classical monocytes **(C)** Clinical Frailty Score **(D)** Aspartate transaminase **(E)** Proportion of natural killer cells **(F)** proportion of B cells **(G)** Soluble CD25 **(H)** Proportion of intermediate monocytes. MFI=median frequency intensity

